# UTILIZATION OF PRECONCEPTION CARE AND ASSOCIATED FACTORS AMONG REPRODUCTIVE AGE WOMEN IN POKHARA METROPOLITAN, KASKI, NEPAL

**DOI:** 10.1101/2024.10.05.24314942

**Authors:** Sita Aryal, Saroj Wagle, Yamuna Marasini, Tulsi Ram Bhandari

**Author notes:** **Corresponding Author Sita Aryal**.

## Abstract

**Background:** Preconception care refers to specific actions taken by an individual or couple to improve their health before becoming pregnant. Preconception care (PCC) is considered primary prevention for the future baby and secondary prevention for prospective mothers. Preconception care utilization in Nepal was very low. Studies on these issues are a newer concept in Nepal and also in Pokhara Metropolitan.

**Objectives:** To assess the utilization of preconception care and associated factors among reproductive-age group women in Pokhara Metropolitan.

**Methods:** A community-based cross-sectional study was conducted from January 2024 to September 2024. The total study participants were 384 reproductive-age women. Data were collected using pre-tested, interviewer-administered questionnaires and analyzed with SPSS after entry into Epi-Data. Descriptive analysis was conducted to show frequencies and percentages, while bivariate analysis was performed using a chi-square test and multivariate analysis was done using binary logistic regression to identify factors associated with the utilization of preconception care. Lastly, odds ratios and 95% confidence intervals were used to evaluate the statistical significance between the dependent and independent variables.

**Results:** Only 0.3% of participants utilize all preconception care components before their last pregnancy. The acceptance level of utilization of the PCC component was 44.5%. The commonest item received by the participants was taking a healthy diet (49.2%), checking weight (43.8%), and blood group screening (53.4%). Age of the respondent AOR=13.89; (CI:2.49-77.33), religion AOR=3.1;CI(1.43-6.95), occupations of respondents AOR=2.58; CI(1.67-3.99), knowledge of PCC AOR=5.12; CI(2.99-8.77), attitude towards PCC AOR=1.92; CI(1.18-3.12), heard about PCC AOR=3.15; CI(1.77-5.59) minutes to reach health facilities AOR=2.4; CI (1.32-4.35) health insurance use before pregnancy AOR=2.68 ;CI(1.71-4.19), and waiting time at health facilities AOR=1.60;CI(1.02-2.50) were identified as a factors associated with utilization of PCC.

**Conclusion:** The utilization of preconception care among the women was very low compared to the recommended services and was influenced by various factors.

Community-based advocacy and awareness are needed for women with low levels of knowledge and attitudes towards preconception care components, as well as those facing challenges such as time to reach health facilities, lack of insurance use before pregnancy, and long waiting times at health facilities. Health promotion strategies focused on preconception care and pre-pregnancy planning may increase utilization rates and improve maternal health.

## Introduction

Preconception care describes specific actions taken by an individual or couple to improve their health before becoming pregnant Preconception care involves provision of biomedical, behavioral and social health interventions to women before conception. Preconception aim to enhance health status and subside behavioural and environmental factors that could lead to adverse outcomes for both maternal and child health^1^.

The World Health Organization recognizes that providing preconception care can help reduce baby and maternal morbidity and mortality The World Health Organization addresses several parts of the PCC package, such as dietary conditions, tobacco use, environmental health, infertility, interpersonal violence, pregnancy timing, genetic determinants sexually transmitted infections, HIV, and mental health ^1^. Additionally, the goals of preconception care include identifying concerns regarding genetic conditions, family history, substance abuse, chronic illnesses, infectious diseases, and proper nutrition and folic acid intake. It also seeks to guide weight management. Vaccinations, family planning, psychosocial issues, domestic abuse, and appropriate housing conditions are among the other things it entails advising on^2^.

Preconception care (PCC) is classified as primary prevention for the future infant and secondary prevention for prospective mother ^3^. Preconception care is important not just for family planning and lowering maternal and neonatal mortality, but it also improves long-term health outcomes for teenage girls, women, and children. Adolescent health, particularly reproductive health, is increasingly recognized as an important component of the healthcare continuum. Preconception care is widely recognized as a technique to improve women’s health through medical and behavioral changes before conception, with the ultimate goal of enhancing pregnancy outcomes^2^.

Reducing maternal mortality is one of the key global health priorities. Sustainable Development Goal 3 (SDG) targets to reduce the Maternal Mortality Ratio (MMR) to less than 70 per 1,00,000 live births and the newborn mortality rate to less than 12 per 1000 live births by 2030^4^. But globally, maternal deaths occur every two minutes which accounts for nearly 800 deaths per day due to preventable pregnancy and childbirth complications. In 2020, total estimated maternal deaths (2,53,000) worldwide Ninety-five percentage of maternal death occurs in low and lower-middle-income countries, accounting for 87% in Sub-Sahara Africa and Southern Asia ^5^. The maternal mortality ratio is 412 per 100,000 live births similarly, the under-five mortality rate is 67 per 1000 live birth and the neonatal mortality rate is 29 per 1000^6^. Similarly, each year, approximately 208 million pregnancies take place out of them, 46% are unintended pregnancy ^7^.

In Nepal, 653 of the 12,976 deaths in 2021 among women aged 15 to 49 were pregnancy-related, with 622 confirmed as maternal deaths ^8^According to studies, induced abortion was responsible for half of all maternal deaths in Nepal, where abortion was illegal until September 2002. Unsafe practices by traditional birth attendants lead to maternal mortality. According to Nepal Demographic Health Survey (2006) revealed that 82% of women gave birth at home. However, by 2022, this ratio had improved to only 19% which indicates that women are still delivering at home (NDHS 2022). Although there is huge progress in maternal and child health but still, the maternal mortality ratio is 151 per one lakh live births, while infant and neonatal mortality rates are 28 and 21 deaths per 1,000 live births, respectively (NPHC report, 2021; NDHS report, 2022).

Most of the complications related to pregnancy and adverse birth outcomes could be prevented through the PCC strategy approach.^9^ Preconception care (PCC) helps to identify and prevent congenital abnormalities such as fetal alcohol spectrum disorders, congenital heart diseases, neural tube defects, and clefts ^10^. The main modifiable risk factors for stillbirth are dietary variables like obesity or undernutrition, maternal age, maternal infection, and non-communicable disorders like diabetes or hypertension^11^. Anemia has been identified as a substantial risk factor for maternal death, increasing the likelihood of maternal mortality during and after childbirth ^11^. Perinatal fatalities are increased by a 50% greater chance among children who are born by a mother under the age of 20 while comparing women aged 20 to 29 ^1^. Violence against girls and women has negative physical, psychological, and reproductive repercussions, including an increased chance of premature delivery and low-birth-weight infants^9^.

The evidence shows that the utilization of PCC is low. Studies conducted in developing countries such as Ethiopia^12^, Nigeria ^13^Bhutan^10^, Nepal^14^ and Sri Lanka ^15^ revealed preconception care of 22.3%, 23.4%, 21.8%, 2% and 27.2% respectively. similarly the study conducted in Los Angeles (29.7%), and Saudi Arabia (29.3%)^16^ shows a slightly higher rate of utilization of PCC.

Factors associated with utilization of preconception care were age, education status of mother, occupation of mother, husband’s education level, husband’s occupation, house wealth status, planned pregnancy, previous ANC visit, previous PNC visit, use, previous adverse pregnancy outcomes, Knowledge on Preconception care, attitude towards PCC, distance from health facilities, waiting time at health facilities, autonomy to maternal health.^2,10,17,18^

In the context of Nepal, a comprehensive national policy addressing preconception care is not in place as the deadline for meeting the Sustainable Development Goal of reaching a maternal mortality ratio of 70 per 100,000 live births, executing an intervention strategy that integrates a continuum-of-care approach through PCC is crucial to minimizing maternal and child mortality.

Despite its important role in improving mother and child health, a large proportion of women are unaware of how their preconception care can influence to reduce the risk of adverse pregnancy outcomes. In practice, there is a lack of checkups or care taken by women before being pregnant and Antenatal care is too late to reduce the harmful effects that a woman’s (and her partner’s) health risks or health problems may have on the fetus during the critical period of organogenesis. Different studies have been done regarding the knowledge and practices on preconception care among reproductive-age women but there is limited research on finding out the factors associated with utilization of preconception care. This study aims to assess the status of preconception care utilization and its associated factors among reproductive-age women of Pokhara Metropolitan.

**Figure 1:**
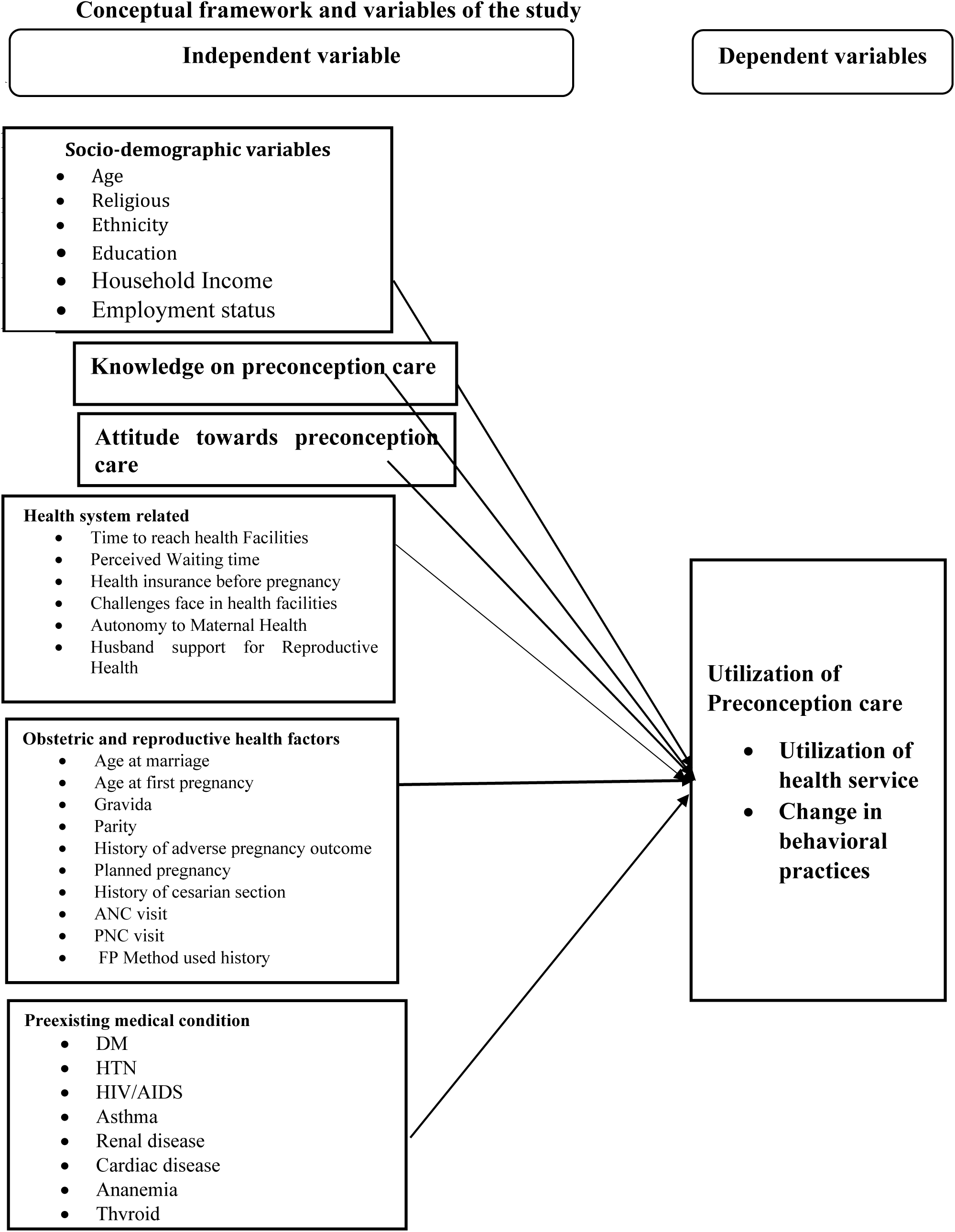
Conceptual Framework.

## Method and materials

### Study design

The study was a community-based cross-sectional study among married women of reproductive age in Pokhara Metropolitan

### Study Setting, participants and sampling procedure

A study was conducted in Pokhara metropolitan from January 2024 to September 2024. A consecutive sampling was carried out among participant. Inclusion criteria was, all Married women of reproductive age having a child under one year of age, women living in selected wards for more than 6 months and above were included in the study. The sample size was calculated using the formula 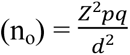 for estimating the required sample with a confidence level of 95% (i.e., Z = 1.96). The allowable error (d) was considered 0.05 (5%). The prevalence of utilizations of preconception care was unknown; thus, the estimated sample size was 384. Data were collected through proportionate sampling method and sample size was determine through consecutive sampling.

### Instrument

The research tool was a structured interview schedule adopted from the study conducted in Ethiopia (Setegn Alie et al., 2022) and designed in the Nepali language. The schedule comprised eight Parts:

Part 1 is Socio-demographic characteristics comprises 10 items Part 2 is knowledge of preconception care comprises 25 items. Each item has Yes and No answers “Yes” counted as 1 and “No” counted as 0. The score of knowledge level ranges from 1 to 25. After scoring, the result of the median values is 18. The score which is less or equal to 18 is categories as poor knowledge and more than 18 as good knowledge. Part C utilization of preconception care which consists of 18 items each items has “Yes” and “No”, answer “Yes” counted as 1 and “No” counted as 0 the score was summed then the total score was ranges from 1 to 18, and the median score is 5. The score which is less or equal to 5 is categorised as low acceptance and more than 5 is regarded as acceptance, Part D attitude towards preconception care consists of 11 items. Likert scale with scores of 1 = “strongly disagree”, 2 = “disagree”, 3 = “neutral”, 4 = "agree,” and 5 = strongly agree” was used. Similarly, for negatively worded items, the score was reversed. The total score was summed up, and the final score ranges from 24 to 51; the median value score is 40. The score less than or equal to 40 is categorised as negative attitude and more than 40 is categorized as positive attitude. Part E pre-existing medical illness, Part F is obstetric and reproductive history consists of 9 items Part G-health facility related 17 items and Part H-wealth of the household consists of 12 items, measured through the International Wealth Index, which measures the economic situation of households based on the possession of consumer durables and housing characteristics. IWI ranges from 0 to 100; a percentile is used to categories the ranges. and categorized as lowest, Second, middle, fourth and highest based on Nepal demography health survey.

### Data Analysis

Descriptive statistics such as frequency, percentage, mean, median and standard deviation were used to present the findings in the form of a table and figures. Bivariate analysis was performed to find out the association between dependent and independent variables and multivariate logistic regression models were analyzed to determine factors associated with the utilization of preconception care. variables with a p-value less than 0.05 in the bivariable logistic regression were included in the multivariable logistic regression model based on the conceptual framework three models of logistic regression were performed. In the first model dependent variable utilization of PCC and Sociodemographic characteristics were assessed, in the second model utilization of PCC and knowledge of PCC, attitude towards PCC were assessed. In the third model, the utilization of PCC and health facilities service-related characteristics were assessed. The multicollinearity or interaction between independent variables were checked using the VIF (Variance Inflation factor) and all the variables less than 2 indicated as no multicollinearity issues. The strength of association between dependent and independent variables was measured using both Unadjusted Odds Ratio (UOR) and Adjusted Odds Ratio (AOR) along with their 95% CI and p-values. Variables with p-values less than 0.05 were considered statistically significant. The goodness of fit of the model was assessed using the Hosmer and Lemeshow test, the p-value more than 0.05 was considered as good for model fitness.

### Ethical consideration

Ethical approval was taken from the Institutional Review Committee (IRC) of Pokhara University (Ref No.194-2080/81), before conducting the study. Approval was also taken from the local authorities, Pokhara Metropolitan Health Section. The purpose of the study was clearly explained to the participants, and both verbal and written consent were obtained from each participant. They were also informed of their right to withdraw from the survey at any time if they felt uncomfortable continuing.

## Results

### Univariate analysis

Socio-demographic characteristics of respondents (n=384)

Majority of the participants were between 20-35 years (88%), Most people are Hindu (90.9%), with a small portion of Buddhists (4.9%), Christians (2.9%), and Muslims (1.3%). Brahman/ Chhetri (42.4%) is the largest group, followed by Janajati, Dalit and religious minorities by 30.7%, 22.1% and 4.7% respectively. Nearly similar respondents lived in nuclear (45.3%) and joint families (46.1%) with only 8.6% in extended families. Family size is also roughly similar, with 49.5% having four or fewer members and 50.5% having more than four members (See Table 1).

**Table 1:** Socio-demographic characteristics.

| Variables | Number | Percent (%) |
| --- | --- | --- |
| Age in years |  |  |
| ≤ 20 | 22 | 5.7 |
| 20-35 | 338 | 88.0 |
| >35 | 24 | 6.3 |
| Mean =28.43±4.94, min=16, max=44 |  |  |
| **Religion** |  |  |
| Hindu | 349 | 90.9 |
| Buddhist | 19 | 4.9 |
| Muslim | 5 | 1.3 |
| Christian | 11 | 2.9 |
| **Ethnicity** |  |  |
| Dalit | 85 | 22.1 |
| Janajati | 118 | 30.7 |
| Brahman/ Chhetri | 163 | 42.4 |
| Religious Minorities | 18 | 4.7 |
| **Family type** |  |  |
| Nuclear | 174 | 45.3 |
| Joint | 177 | 46.1 |
| Extended | 33 | 8.6 |
| **Family size** |  |  |
| ≤ 4 | 190 | 49.5 |
| >4 | 194 | 50.5 |

### Socio-economic characteristics of respondents (n=384)

**Table 2:** Socio-economic Characteristics.

| **Variables** | **Number** | **Percent (%)** |
| --- | --- | --- |
| **Education level** |  |  |
| Illiterate | 5 | 1.3 |
| Informal Education | 7 | 1.8 |
| Basic | 76 | 19.8 |
| Secondary | 199 | 51.8 |
| Graduate and above | 97 | 25.3 |
| **Husband Education Level** |  |  |
| Illiterate | 9 | 2.3 |
| Informal | 15 | 3.9 |
| Basic | 76 | 19.8 |
| Secondary | 178 | 46.4 |
| Graduate and above | 106 | 27.6 |
| **Respondent Occupation** |  |  |
| Housemaker | 209 | 54.4 |
| Farmer | 18 | 4.7 |
| Business | 56 | 14.6 |
| Government Employee | 22 | 5.7 |
| Unemployed | 22 | 5.7 |
| Private Employer | 43 | 11.2 |
| Daily worker | 10 | 2.6 |
| Foreign services | 4 | 1.0 |
| **Husband Occupation** |  |  |
| Farmer | 8 | 2.1 |
| Business | 92 | 24.0 |
| Government Employee | 29 | 7.6 |
| Unemployed | 4 | 1.0 |
| Private Employer | 75 | 19.5 |
| Daily worker | 71 | 18.5 |
| Foreign services | 105 | 27.3 |
| **Wealth Index** |  |  |
| Lowest | 73 | 19.0 |
| Second | 86 | 22.4 |
| Middle | 83 | 21.6 |
| Fourth | 65 | 16.9 |
| Highest | 77 | 20.1 |

IWI median=64.48, Min=21, Max=100

The majority of the respondents 51%have their secondary education, with their husbands having secondary level education comprising (46.4%). Regarding respondents’ occupation, most respondents are housemakers (54.4%) while for husbands’ occupation, most of them are foreign services (27.3%). regarding the household index, the second quintile has the highest proportion at (22.4%). (Table-2).

### Heard about Preconception Care and Source of Information

**Table 3:** Heard about Preconception Care and Source of Information.

| **Variables** | **Number** | **Percent (%)** |
| --- | --- | --- |
| **Heard about Preconception care** |  |  |
| Yes | 226 | 58.9 |
| No | 158 | 41.1 |
| **Sources of information (multiple responses, n=513)** |  |  |
| Health worker | 132 | 58.4 |
| School | 68 | 30.1 |
| Neighbors | 44 | 19.5 |
| Mass media | 145 | 64.2 |
| family and friends | 124 | 54.9 |

### Knowledge, attitude and utilization level of respondents

Less than half (47.7%) of respondents have good knowledge while 46.1% exhibit a negative attitude towards preconception care furthermore, only (44.5%) of the respondents utilize preconception care at the acceptance level (Table 4,5,6).

**Table 4:** Preconception Knowledge Attitude Level of Participants.

| Level of knowledge | Frequency | Percent (%) |
| --- | --- | --- |
| Poor Knowledge | 201 | 52.3 |
| Good Knowledge | 183 | 47.7 |
| Median=18, Min=1, Max=25 |  |  |

**Table 5:** Preconception Knowledge Attitude Level of Participants.

| Level of attitude |  |  |
| --- | --- | --- |
| Negative Attitude | 207 | 53.9 |
| Positive Attitude | 177 | 46.1 |
| Median=40, Min=24, Max=51 |  |  |

**Table 6:**
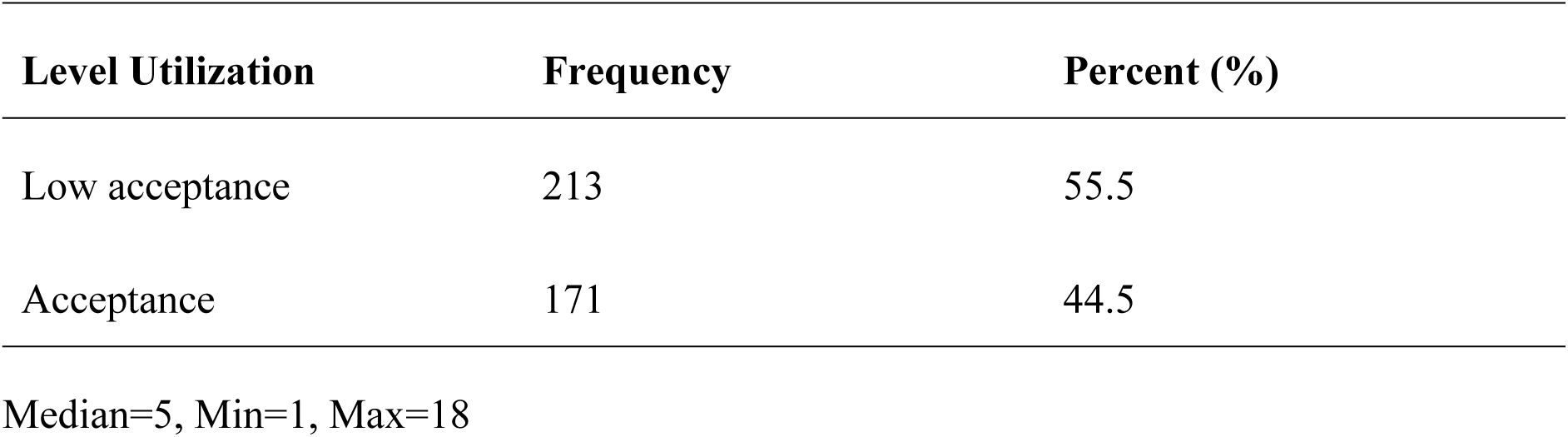
Utilization of Preconception Care Component.

### Preconception Care Components Utilization

Most of the respondents did not take folic acid (64.1%) or iron supplements (91.7%), Similarly the highest proportion did not screen for HIV/AIDS (72.9%), STIs (72.4%), diabetic mellitus (66.7%), or hypertension (53.4%). Furthermore, 64.8% did not consult anyone for advice about becoming pregnant (Table 7).

**Table 7:** Preconception Care Component Utilization.

| Variable | Frequency | Percent (%) |
| --- | --- | --- |
| **Take folic acid** |  |  |
| Yes | 138 | 35.9 |
| No | 246 | 64.1 |
| **Take iron** |  |  |
| Yes | 32 | 8.3 |
| No | 352 | 91.7 |
| **Take a healthy diet** |  |  |
| Yes | 189 | 49.2 |
| No | 195 | 50.8 |
| **Take extra meal** |  |  |
| Yes | 35 | 9.1 |
| No | 349 | 90.9 |
| **Check weight** |  |  |
| Yes | 168 | 43.8 |
| No | 216 | 56.3 |
| **Screen for HIV/AIDS** |  |  |
| Yes | 104 | 27.1 |
| No | 280 | 72.9 |
| **Screen for STI** |  |  |
| Yes | 106 | 27.6 |
| No | 278 | 72.4 |
| **Screen for Diabetic mellitus** |  |  |
| Yes | 128 | 33.3 |
| No | 256 | 66.7 |
| **Screen for hypertension** |  |  |
| Yes | 179 | 46.6 |
| No | 205 | 53.4 |
| **Screen for anaemia** |  |  |
| Yes | 153 | 39.8 |
| No | 231 | 60.2 |
| **Screen for blood group** |  |  |
| Yes | 205 | 53.4 |
| No | 179 | 46.6 |
| **Screen for Hepatitis B** |  |  |
| Yes | 40 | 10.4 |
| No | 344 | 89.6 |
| **Take tetanus vaccine** |  |  |
| Yes | 21 | 5.5 |
| No | 363 | 94.5 |
| **Ever smoked** |  |  |
| Yes | 12 | 3.1 |
| No | 372 | 96.9 |
| **Drinking alcohol** |  |  |
| Yes | 23 | 6 |
| No | 361 | 94 |
| **Consult anyone for advice about becoming pregnant** |  |  |
| Yes | 135 | 35.2 |
| No | 249 | 64.8 |
| **Husband has screened for any disease** |  |  |
| Yes | 82 | 21.4 |
| No | 302 | 78.6 |
| **Stopped or removed family planning method** |  |  |
| Yes | 83 | 21.6 |
| No | 301 | 78.4 |

### Pre-existing medical illness

Of the total 384 respondents, 46 (12%) of them experienced any choric diseases in their life. The commonest diseases were thyroid 16 (34.8%), Hypertension 9 (19.6%), Diabetic Mellitus 7 (15.2%) and the least common diseases were anemia, cardiac diseases and renal disease i.e.2(4.3%) for each (Table 8).

**Table 8:** Pre-existing medical illness.

| **Variables** | **Frequency** | **Percent (%)** |
| --- | --- | --- |
| **Medical illness** |  |  |
| Yes | 46 | 12 |
| No | 338 | 88 |
| **Tye of medical illnesses (Multiple responses, n=48) \*** |  |  |
| Epilepsy | 3 | 6.5 |
| Hypertension | 9 | 19.6 |
| Diabetes mellitus | 7 | 15.2 |
| Anemia | 2 | 4.3 |
| Asthma | 4 | 8.7 |
| Cardiac problem | 2 | 4.3 |
| Renal disease | 2 | 4.3 |
| Thyroid | 16 | 34.8 |
| other medical illness | 3 | 6.5 |

### Obstetric and Reproductive Health Characteristics

The third quartile (75.8%) of the respondents had their first pregnancy after age 20 and the majority (53.6%) had only one delivery. Planned pregnancy comprised (64.4%) and (28.6%) had a history of adverse pregnancy outcomes with spontaneous abortion being the most common adverse outcome (54.5%). Similarly, more than one-fourth (32.6%) had a history of cesarean section, and 65.9% had never used contraceptives. Nearly all the respondents (93.7%) had ANC visits for their last pregnancy, with (93.7%) having four or more visits (Table 9).

**Table 9:** Obstetric and Reproductive Health Characteristics.

| **Variables** | **Frequency** | **Percent (%)** |
| --- | --- | --- |
| **Age at first pregnancy** |  |  |
| ≤ 20 | 93 | 24.2 |
| > 20 | 291 | 75.8 |
| Mean=23, Min=13, Max=41 |  |  |
| **Gravida** |  |  |
| only 1 pregnancy | 157 | 40.9 |
| 2-4 pregnancy | 222 | 57.8 |
| ≥ 5 | 5 | 1.3 |
| **Parity** |  |  |
| Primipara (one deliveries) | 206 | 53.6 |
| Multi Parous (more than or equal to 2 deliveries) | 178 | 46.4 |
| **Planning for pregnancy** |  |  |
| Yes | 259 | 67.4 |
| No | 125 | 32.6 |
| **History of adverse pregnancy outcome** |  |  |
| Yes | 110 | 28.6 |
| No | 274 | 71.4 |
| **Type of adverse pregnancy outcome (multiple responses, n=110) \*** |  |  |
| Spontaneous abortion history | 54 | 54.5 |
| stillbirth history | 7 | 7.1 |
| preterm birth history | 26 | 26.3 |
| congenital abnormality history | 11 | 11.1 |
| neonatal death history | 6 | 6.1 |
| low birth weight baby weight | 38 | 38.4 |
| **Cesarean section History** |  |  |
| Yes | 125 | 32.6 |
| No | 259 | 67.4 |
| **History of contraceptive use** |  |  |
| Yes | 131 | 34.1 |
| No | 253 | 65.9 |
| **Previous use of family planning** |  |  |
| IUCD | 2 | 1.5 |
| Implant | 13 | 9.9 |
| Depo | 52 | 39.7 |
| Oral Contraceptive | 15 | 11.5 |
| Condom | 49 | 37.4 |
| **ANC visit for the index pregnancy** |  |  |
| Yes | 381 | 99.2 |
| No | 3 | 0.8 |
| **Previous ANC visit for nearest pregnancy (n=382)** |  |  |
| 1-2 | 11 | 2.9 |
| 3 | 13 | 3.4 |
| ≥ 4 | 358 | 93.7 |
| **PNC visit for index pregnancy** |  |  |
| Yes | 344 | 89.6 |
| No | 40 | 10.4 |
| **Times of PNC visit(n=146)** |  |  |
| 1-2 | 310 | 89.6 |
| ≥ 3 | 36 | 10.4 |

*Note – \*-Multiple responses (the total sum of the percentage is more than 100% because of multiple* *responses).*

### Health service-related characteristics

**Table 10:** Health services-related characteristics.

| **Variables** | **Number (n=384)** | **Percent %)** |
| --- | --- | --- |
| **Minutes to reach Health facilities** |  |  |
| ≤ 30 minutes | 316 | 82.3 |
| >30 minutes | 68 | 17.7 |
| **Distance from health facilities** |  |  |
| ≤ 2 km | 148 | 38.5 |
| > 2 km | 236 | 61.5 |
| **means of transport** |  |  |
| Public Vehicle | 154 | 40.1 |
| Private Vehicle | 128 | 33.3 |
| Walk | 102 | 26.6 |
| **Visit health facilities for a healthy pregnancy** |  |  |
| Yes | 125 | 32.6 |
| No | 259 | 67.4 |
| **Pay to get services(n=125)** |  |  |
| Yes | 97 | 77.6 |
| No | 28 | 22.4 |
| **Affordability of payment(n=97)** |  |  |
| Expensive | 35 | 36.5 |
| Cheap | 52 | 53.6 |
| Free | 10 | 10.3 |
| **Participating in preconception counselling(n=125)** |  |  |
| Yes | 96 | 77.4 |
| No | 28 | 22.6 |
| **Type of counselling (multiple response (n=96)** |  |  |
| Maintaining optimal weight control | 43 | 44.8 |
| maintaining a regular exercise program | 54 | 56.3 |
| ceasing tobacco, alcohol, and drug use | 58 | 60.4 |
| Follows the prescription of the health worker | 90 | 93.8 |
| Take healthy diet | 96 | 100 |
| **Challenges faced by respondents in Health facilities(n=124)** |  |  |
| Yes | 59 | 47.2 |
| No | 66 | 52.8 |
| **Type of challenges (multiple responses) (n=79)** |  |  |
| Lack of privacy | 9 | 15.3 |
| Lack of service integrity | 5 | 8.5 |
| Shortage of logistic supply | 13 | 22.0 |
| Long waiting hour | 52 | 88.1 |
| **Availability of adequate laboratory services** |  |  |
| Yes | 303 | 78.9 |
| No | 62 | 16.1 |
| Don't know | 19 | 4.9 |
| **Availability of medicine** |  |  |
| Yes | 360 | 93.8 |
| No | 20 | 5.2 |
| Don't Know | 4 | 1 |
| **Using Health insurance** |  |  |
| Yes | 151 | 39.3 |
| No | 233 | 60.7 |
| **Autonomy to maternal health care services** |  |  |
| Self | 228 | 59.4 |
| Husband | 25 | 6.5 |
| Both | 129 | 33.6 |
| Family member | 2 | 0.5 |
| **Receive health care access assistance (Multiple response (n=717))** |  |  |
| Husband | 282 | 73.4 |
| Relatives | 48 | 12.5 |
| Family | 186 | 48.4 |
| Neighbours | 58 | 15.1 |
| Self | 143 | 37.2 |
| **Perceived waiting time to get services** |  |  |
| ≤ 60 minutes | 236 | 61.5 |
| > 60 minutes | 148 | 38.5 |

The result revealed that 82.3% of the respondents reached health facilities within 30 minutes and 61.5% lived within ≤ 2 km distance from health facilities. Public vehicle is a common vehicle used by respondents (40.1%). Only 32.6% visited health facilities for healthy pregnancies of those who visited, 77.6% needed to pay for their services and out of them 36.5% finds expensive and 77.4% took part in preconception counselling. The challenges face by 47.2%respondents in health facilities and most commonly due to long waiting hour. However, 78.9% of participants revealed that 78.9% had adequate laboratory services and 93.8% had access to medicine. Only 39.3% use health insurance and autonomy to maternal health services was mainly self-directed (59.4%). lastly, 61.5% perceived a waiting time of ≤ 60 to get services (Table-10).

### Bivariate analysis

### Association of Utilization of Preconception Care and Sociodemographic Characteristics

The major findings indicate the age group (χ² = 13.279, p-value < 0.001), religion (χ² = 6.99, p-value = 0.008), ethnicity (χ² = 28.965, p-value < 0.001), and family size (χ² = 3.895, p-value = 0.048), were significantly associated with the utilization of preconception care (Table 11).

**Table 11:** Association of Utilization of Preconception Care and Sociodemographic Characteristics.

| Variable | Utilization of<br>Preconception care |  | Chi-square<br>Value | df | p-value |
| --- | --- | --- | --- | --- | --- |
|  | Low<br>acceptance<br>n (%)<br>213(55.5%) | Acceptance<br>n (%)<br>171(44.5%) |  |  |  |
| Age |  |  |  |  |  |
| < 20 Yrs | 20 (90.9) | 2 (9.1) | 13.279 | 2 | <0.001* |
| 20-35 Yrs | 183 (54.1) | 155 (45.9) |  |  |  |
| > 35 Yrs | 10 (41.7) | 14 (58.3) |  |  |  |
| Religion |  |  |  |  |  |
| Hindu | 201 (57.6) | 148 (42.4) | 6.99 | 1 | 0.008 |
| Other than Hindu | 12 (34.3) | 23 (65.7) |  |  |  |
| Ethnicity |  |  |  |  |  |
| Dalit | 66 (77.6) | 19 (22.4) | 28.965 | 3 | <0.001* |
| Janajati | 69 (58.5) | 49 (41.5) |  |  |  |
| Brahman/Chettri | 69 (42.3) | 94 (57.7) |  |  |  |
| Religious<br>Minorities | 9 (50) | 9 (50) |  |  |  |
| Family size |  |  |  |  |  |
| ≤ 4 | 115 (60.5) | 75 (39.5) | 3.895 | 1 | 0.048 |
| >4 | 98 (50.5) | 96 (49.5) |  |  |  |

### Association of Preconception Care Utilization and Socio-economic Characteristics

There is an association between the utilization of preconception care and the respondent’s socio-economic characteristics. Respondent education level (χ² = 48.541, p-value < 0.001), husband’s education level (χ² = 47.395, p-value < 0.001), respondent occupation (χ² = 27.158, p-value < 0.001), husband’s occupation (χ² = 14.764, p-value = 0.002) and the international wealth index (χ² = 36.851, p-value < 0.001) displayed a significant association (Table 12).

**Table 12:**
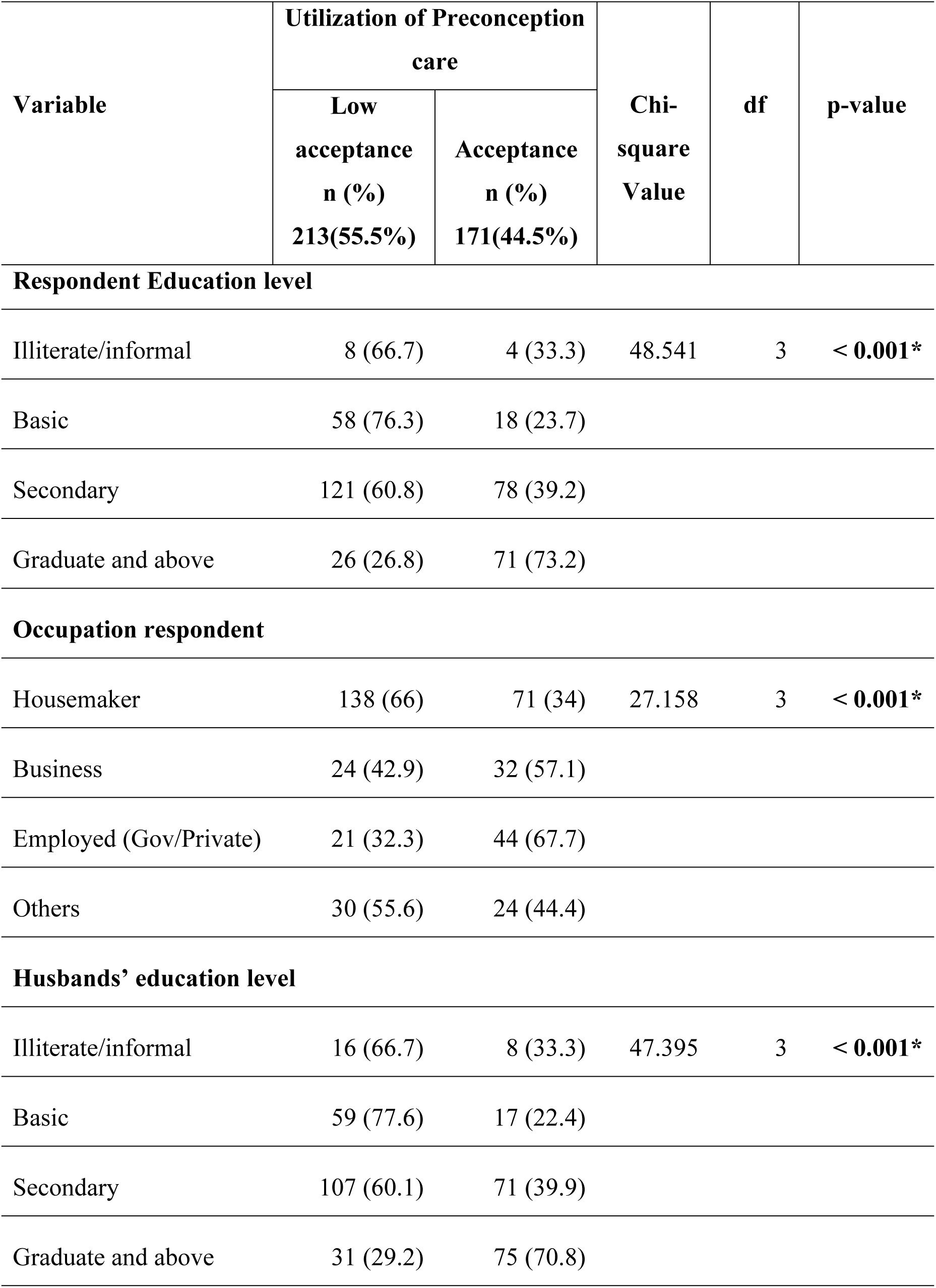
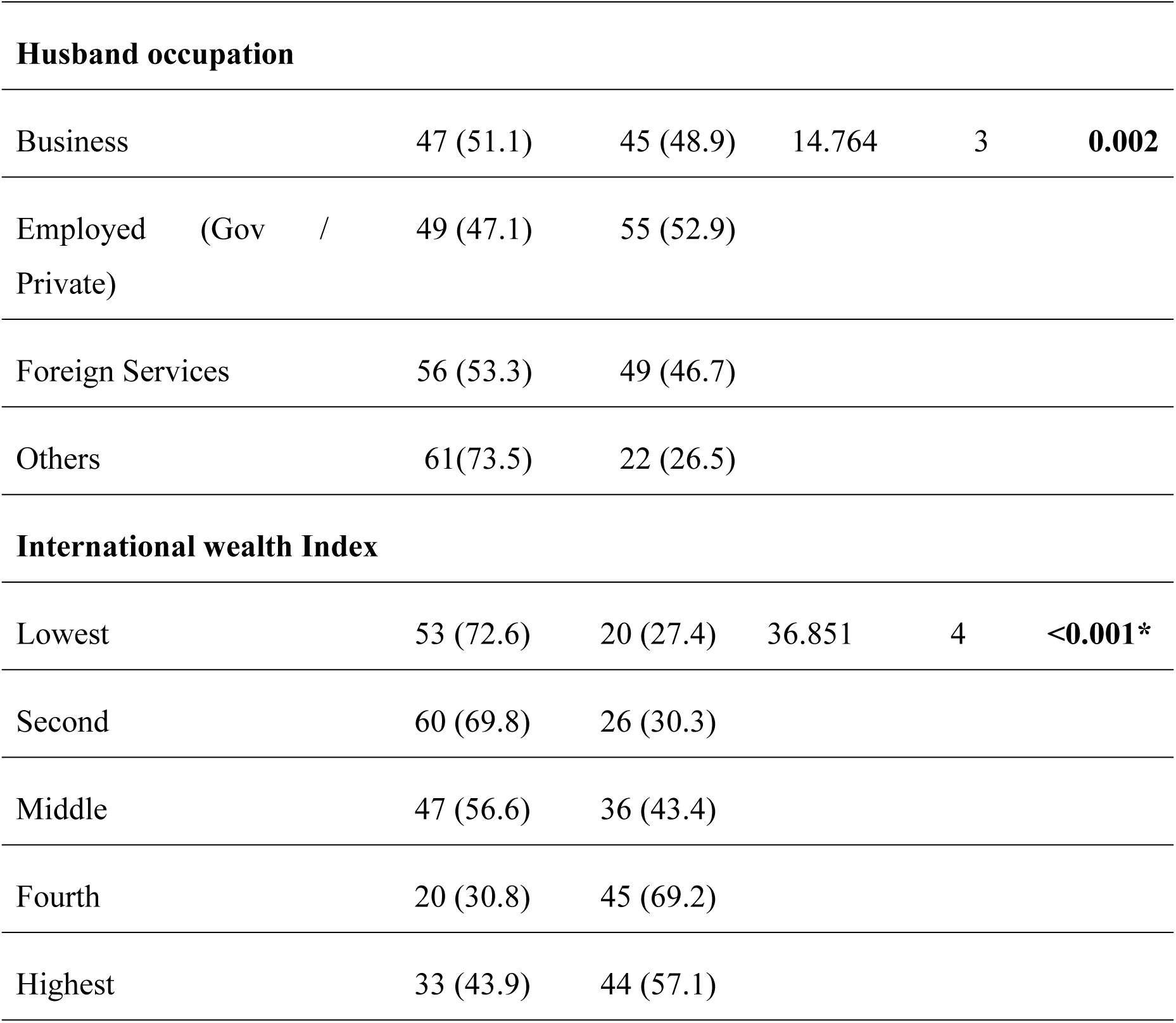
Association of Preconception Care Utilization and Socio-economic Characteristics.

### Association between Knowledge, attitude and heard about PCC with Preconception Care Utilization

The utilization of preconception care shows significant associations with respondents’ knowledge (χ² = 99.448, p-value < 0.001), attitude (χ² = 17.278, p-value < 0.001) and prior awareness of preconception care (PCC) (χ² = 74.473, p-value < 0.001) (Table 13).

**Table 13:** Association between Knowledge, attitude and heard about PCC with Preconception Care Utilization.

| Variable | Utilization of Preconception care |  | Chi-square Value | Df | p-value |
| --- | --- | --- | --- | --- | --- |
|  | Low acceptance<br>n (%)<br>213(55.5%) | Acceptance<br>n (%)<br>171(44.5%) |  |  |  |
| Knowledge on PCC |  |  |  |  |  |
| Poor Knowledge | 160 (79.6) | 41 (20.4) | 99.448 | 1 | <0.001* |
| Good Knowledge | 53 (29) | 130 (71) |  |  |  |
| Attitude on PCC |  |  |  |  |  |
| Negative Attitude | 135 (65.2) | 72 (34.8) | 17.278 | 1 | <0.001* |
| Positive Attitude | 78 (44.1) | 99 (55.9) |  |  |  |
| Heard about PCC before pregnancy |  |  |  |  |  |
| NO | 129 (81.6) | 29 (18.4) | 74.473 | 1 | < 0.001* |
| Yes | 84 (37.2) | 142 (62.8) |  |  |  |

### Association between utilization of PCC with history of obstetrics and reproductive health

The above table shows that there are significant associations with several variables. Age at first pregnancy (χ² = 15.477, p-value < 0.001), planning for pregnancy (χ² = 64.549, p-value < 0.001) and history of adverse health (χ² = 11.630, p-value < 0.001) was significantly associated with the utilization of PCC. However, there is no association with different variables such as include the number of live births (χ² = 2.897, p-value = 0.089), PNC checkup during the last pregnancy (χ² = 3.817, p-value = 0.051), contraceptive use history (χ² = 2.083, p-value = 0.14), and history of Cesarean section (χ² = 3.337, p-value = 0.06) (Table 14).

**Table 14:** Association between utilization of PCC with history of obstetrics and reproductive health.

| Variables | Utilization of Preconception care |  | Chi-square Value | Df | p-value |
| --- | --- | --- | --- | --- | --- |
|  | Low acceptance<br>n (%) 213 (55.5%) | Acceptance<br>n (%)171 (44.5%) |  |  |  |
| Age at first pregnancy |  |  |  |  |  |
| ≤ 20 yrs | 68 (73.1) | 25 (26.9) | 15.477 | 1 | <0.001 |
| > 20 yrs | 145 (49.8) | 146 (50.2) |  |  |  |
| **No of live birth** |  |  |  |  |  |
| Primipara | 106 (51.5) | 100 (48.5) | 2.897 | 1 | 0.089 |
| Multipara | 107 (60.1) | 71 (39.9) |  |  |  |
| **Planned for pregnancy** |  |  |  |  |  |
| No | 106 (84.8) | 19 (15.2) | 64.549 | 1 | < 0.001 |
| Yes | 107 (41.3) | 152 (58.7) |  |  |  |
| **History of adverse health outcome** |  |  |  |  |  |
| No | 167 (60.9) | 107 (39.1) | 11.630 | 1 | < 0.001 |
| Yes | 46 (41.8) | 64 (58.2) |  |  |  |
| **PNC checkup last pregnancy** |  |  |  |  |  |
| No | 28 (70) | 12 (30) | 3.817 | 1 | 0.051 |
| Yes | 185 (53.8) | 159 (46.2) |  |  |  |
| **Contraceptive use history** |  |  |  |  |  |
| No | 147 (58.1) | 106 (41.9) | 2.083 | 1 | 0.14 |
| Yes | 66 (50.4) | 65 (49.6) |  |  |  |
| **History of Cesarean section** |  |  |  |  |  |
| No | 152 (58.7) | 107 (41.3) | 3.337 | 1 | 0.06 |
| Yes | 61 (48.8) | 64 (51.2) |  |  |  |

### Association between utilization of PCC and Health service-related characteristics

There is a significant association between the health service-related variables and utilization of PCC such as time to reach health facilities (χ² = 6.232, p-value = 0.01), health insurance use before pregnancy (χ² = 27.084, p-value < 0.001), autonomy to maternal health (χ² = 8.690, p-value = 0.013), and waiting time at health facilities (χ² = 10.140, p-value< 0.001). However, the distance from health facilities did not show a significant association (χ² = 0.426, p-value = 0.51) with the utilization of preconception care (Table 15).

**Table 15:** Association between utilization of PCC and Health service-related characteristics.

| Variable | Utilization of Preconception<br>care |  | Chi-square<br>Value | Df | p-value |
| --- | --- | --- | --- | --- | --- |
|  | Low | Acceptance |  |  |  |
|  | acceptance | Acceptance |  |  |  |
|  | n (%) | n (%) |  |  |  |
|  | 213(55.5%) | 171(44.5%) |  |  |  |
| **Minutes to reach health facilities** |  |  |  |  |  |
| ≤ 30 minutes | 166 (52.5) | 150 (47.5) | 6.232 | 1 | **0.01** |
| >30 minutes | 47 (69.1) | 21(30.9) |  |  |  |
| Distance from health facilities |  |  |  |  |  |
| ≤2 Km | 79 (53.4) | 69 (46.6) | .426 | 1 | 0.51 |
| > 2 Km | 134 (56.8) | 102 (43.2) |  |  |  |
| **Health insurance use before becoming pregnant** |  |  |  |  |  |
| No | 154 (66.1) | 79 (33.9) | 27.084 | 1 | **<0.001** |
| Yes | 59 (39.1) | 92 (60.9) |  |  |  |
| **Autonomy to Maternal Health** |  |  |  |  |  |
| Self | 114 (50) | 114 (50) | 8.690 | 2 | **0.013** |
| Husband | 19 (76) | 6 (24) |  |  |  |
| Both | 80 (61.1) | 51 (38.9) |  |  |  |
| **Waiting time at health facilities** |  |  |  |  |  |
| ≤ 60 min | 146 (61.9) | 90 (38.1) | 10.140 | 1 | **< 0.001** |
| > 60 min | 67 (45.3) | 81(54.7) |  |  |  |

### 4.3. Multivariate Analysis

Bivariate analysis was performed to find out the association between dependent and independent variables and multivariate logistic regression models were analyzed to determine factors associated with the utilization of preconception care. variables with a p-value less than 0.05 in the bivariable logistic regression were included in the multivariable logistic regression model Based on the conceptual framework three models of logistic regression were performed. In the first model dependent variable utilization of PCC and Sociodemographic characteristics were assessed, in the second model utilization of PCC and knowledge and attitude towards PCC, in the third model utilization of PCC and health service-related characteristics were assessed. Three models were developed, in the first model is age of respondents, Religion, family size, and occupation of the respondent, were assessed, In the second model, knowledge of PCC, attitude towards PCC and heard about were assessed, similarly, in the third model health service-related characteristics i.e., health insurance before pregnancy, minutes to reach health facilities, waiting time at health facilities, autonomy to maternal health were analyzed with dependent variables.

### Relationship between sociodemographic characteristics and Utilization of Preconception care

Respondents aged 20-35 years are 9 times more likely to use preconception care services as compared to those who are under 20 years AOR=9.0; CI (1.99-40.71) Similarly, respondents aged more than 35 years are nearly 14 times more likely to utilize PCC as compared to under 20 years AOR=13.89; CI (2.49-77.33), this show as the age increases the utilization of PCC also increases. participants who follow a religion other than Hinduism have 3.15 times more chances to use PCC as compared to Hindus AOR=3.15, CI (1.43-6.95) Participants engaged in other occupations besides housemaker are 2.58 times more likely to utilize PCC compared to housemaker AOR=2.58; CI (1.67-3.99). The Nagelkerke R square in sociodemographic characteristics is 0.148, which indicates 14.8%of the variance in the utilization of PCC (Table 16)

**Table 16:**
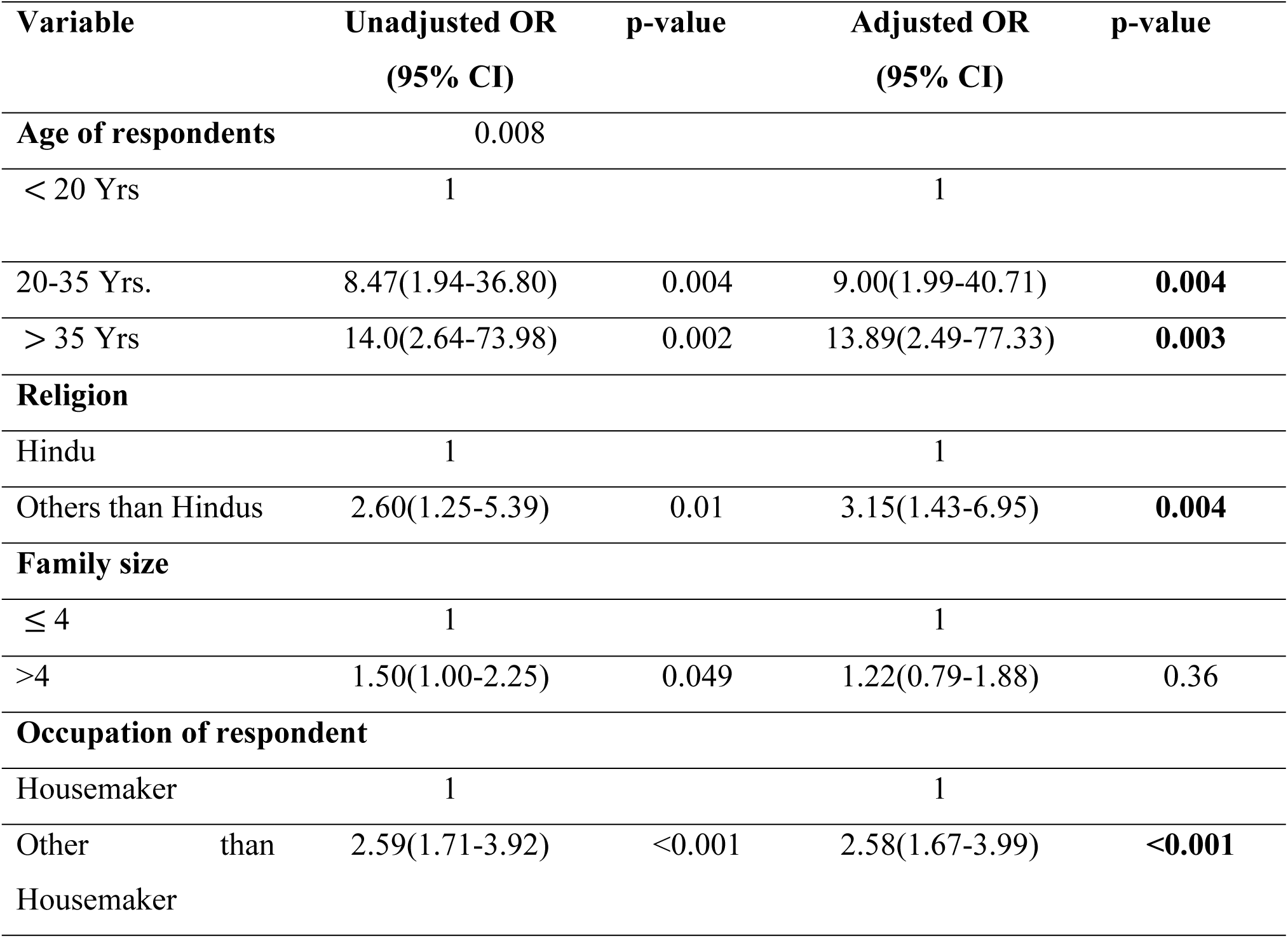
Relationship between sociodemographic characteristics and Utilization of preconception care.

### Relationship between sociodemographic characteristics and Utilization of Preconception care

Result revealed that the respondents who have good knowledge of PCC were more than 5 times more likely to use PCC services as compared to those participants who had poor knowledge AOR=5.12; CI (2.99-8.77). Similarly, respondents who had a positive attitude towards PCC were nearly 2 times more likely to use PCC services as compared to those participants who had a negative attitude AOR=1.92; CI (1.18-3.12) furthermore, the findings underscore respondents who had heard about PCC were more than 3 times more likely to use PCC services while comparing with those who had not heard about PCC AOR=3.15; CI (1.77-5.59). The Nagelkerke R Square of this model is 0.377 which indicates 37.7% variability in utilization of PCC (Table 17).

**Table 17:**
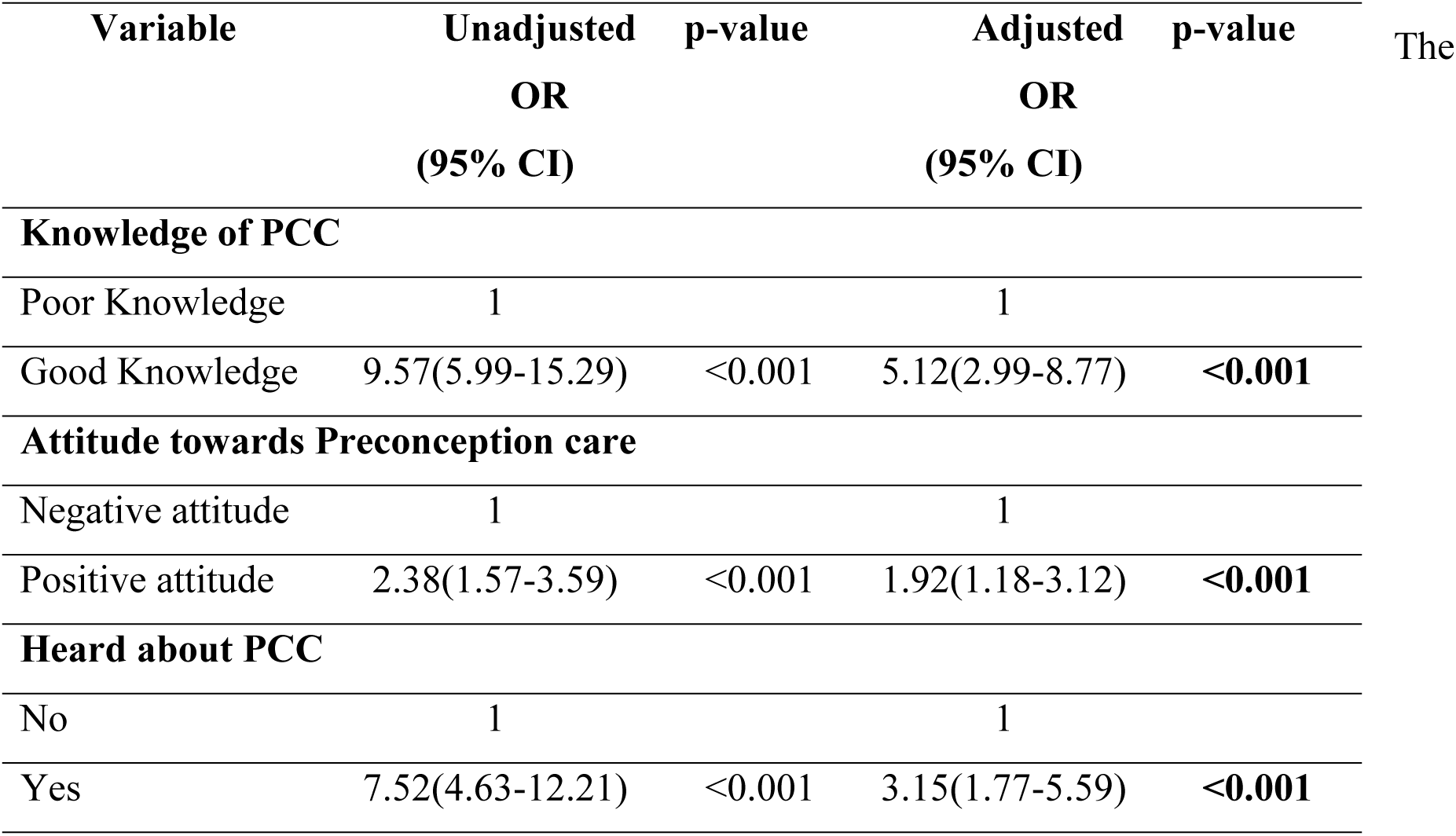
Relationship between knowledge and attitude towards PCC and Utilization of Preconception.

### Relationship between Health Services related characteristics and utilization of PCC

Respondents who reach health facilities within 30 minutes are 2.40 times more likely to utilize PCC compared to than who take more than 30 minutes. Similarly, participants who had health insurance before pregnancy are 2.68 times more likely to use PCC compared to those without health insurance. Respondents who wait more than 60 minutes at health facilities are 1.60 times more likely to utilize PCC compared to those who wait 60 minutes or less. Respondents who have autonomy in their maternal health decisions are 1.45 times more likely to utilize PCC compared to those where decisions are made jointly by both husband and wife, this result is not statistically significant after adjustment. The Nagelkerke R Square of this model is 0.14 which indicates 14%variability in utilization of PCC (Table 18).

**Table 18:** Relationship between Health Services related characteristics and utilization of PCC.

| Variable | Unadjusted<br>OR | p-value | Adjusted<br>OR | p-value |
| --- | --- | --- | --- | --- |

|  | (95% CI) |  | (95% CI) |  |
| --- | --- | --- | --- | --- |
| **Minutes to reach health facilities** |  |  |  |  |
| ≤30 minutes | 2.02(1.15-3.54) | 0.014 | 2.40(1.32-4.35) | **0.004** |
| > 30 minutes | 1 |  | 1 |  |
| **Health Insurance use before becoming pregnant** |  |  |  |  |
| No | 1 |  | 1 |  |
| Yes | 3.04(1.98-4.64) | <0.001 | 2.68(1.71-4.19) | **<0.001** |
| **Autonomy of Maternal Health** |  |  |  |  |
| Self | 1.73(1.14-2.63) | 0.009 | 1.45(0.93-2.27) | 0.09 |
| Both (Husband and wife) | 1 |  | 1 |  |
| **Waiting time at health facilities** |  |  |  |  |
| ≤60 min | 1 |  | 1 |  |
| > 60 min | 1.96(1.29-2.97) | 0.002 | 1.60(1.02-2.50) | **0.04** |

## DISCUSSION

The current study aims to assess preconception care component utilization and its associated factors among reproductive-age women of Pokhara Metropolitan, Nepal.

This study revealed that only 1 (0.3%) of the participants received all 18 preconception care components before they underwent their last pregnancy. The possible explanation for these results might be due to the lack of awareness and less importance given to services of preconception care and mainly attention is given after conception. The acceptance level of preconception care components was 44.5% and the remaining 55% had a low acceptance level of utilization. This study is allied with the prior study conducted in Bheerkot, which indicated a 47.9% of good PCC practices^8^. However, a study in Bhutan demonstrated 21% Good Practice of PCC which is half of the findings of the current research.^10^. Similarly study conducted in Nigeria 24.1%,^19^ Ethiopia (22.3%)^12^ show the lower utilization of PCC. The justification for this may be due to various perceptions of maternal health care in different cultures and variations in the socio-demographic characteristics of the study participants.

Furthermore, this study showed the most common component utilized by women is avoiding/cessation of cigarettes, the women who never smoked in their lives (96.9%) out of those who smoked cigarettes 66.7% stopped or reduced smoking before conception similarly study shows low cessation of smoking in (30.28%) China, Nigeria (27.2%)^19,20^. Taking a healthy diet before conception is high in Bheerkot (90.1%) and Guji Ethiopia (85.7%) and but less practice (49.2%) is seen in this study. Only 35.9% take folic acid which is in lined with study conducted in Bhutan (25%) and much lower in Kathmandu (4%) and in Ethiopia (13.2%) and higher intake in Sri Lanka (43.6%). Taking consulting for a healthy pregnancy is 35.2% in this study which is higher than the study done in Bhutan, Nigeria, Ethiopia ^10,19^. Screening for STI is low (27.6%) in this study which is less than the study conducted in Guji Ethiopia (87.1%), Bhutan (39.7%) and similar to Osun, Nigeria (29.9%). Family planning method removal/stop is 35.9% in this study which is aligned with the study in Bhutan (37.7%)and higher than in Ethiopia (17.9%).^10,18^.

The planned pregnancy in this study was 67.4%, which is higher than the study conducted in Sri Lanka (62%) and South West Ethiopia (44.8%) and lower than the study conducted in Nigeria (77.7%) ^13,15,18^ The findings show there is a significant association between planned pregnancy and utilization of PCC (χ² = 64.549, p-value < 0.001) the result was aligned with a study conducted in Nigeria, Adet, Northwestern Ethiopia^2,13^ the justification for this result might be due to effective family planning interventions are in place. Mothers who had experienced adverse pregnancy outcomes showed statistically significant associations with the utilization of PCC (χ² = 11.630, p-value < 0.001). Similar findings were seen in Southern Ethiopia and Northern Ethiopia ^6^This result might be due to mothers who experience adverse pregnancy outcomes wanting to use PCC services to reduce potential risk for future pregnancy.

In this study, respondents aged 20-35 years are 9 times more likely to use preconception care services as compared to those who are under 20 years AOR=9.0; CI (1.99-40.71) Similarly, respondents aged more than 35 years are nearly 14 times more likely to utilize PCC as compared to under 20 years AOR=13.89; CI (2.49-77.33), this study is allied with the study conducted in Malaysia shows more than two times and in Nigeria 1.3 times utilization of PCC services as compared to their counterparts.^13,21^ this shows as age increases the utilization of PCC also increases. This result might be due to increased health concerns and higher chances of pregnancy complications among older age groups which encourage them to seek PCC services. In this study participants who follow a religion other than Hinduism have 3.15 times more chances to use PCC as compared to Hindus. This may be due to socio-cultural belief or practices regarding preconception care. This study found that participants engaged in other occupations besides housemaker are 2.58 times more likely to utilize PCC compared to housemaker but in contrast, the study conducted in Malaysia, and Nigeria shows women who are not working utilize PCC services ^13,21^ the reason for that was working women are more exposed to the information, and meeting with different people enhances the knowledge seeking for utilization of preconception care.

In this study respondents who had good knowledge of PCC were more than 5 times more likely to use PCC services as compared to those participants who had poor knowledge AOR=5.12; CI (2.99-8.77). These findings is consistent with the study conducted in West Guji Ethiopia and Southern Ethiopia which reported that individuals with poor knowledge of PCC had 47% and 82% reduced the chances of utilizing PCC compared with those with good knowledge.^6,12^. This might be because having good knowledge increases the importance of preconception care which motivates them to utilize the PCC service. This study revealed that the respondents who had a positive attitude towards PCC were nearly 2 times more likely to use PCC services as compared to those with a negative attitude AOR=1.92; CI (1.18-3.12). This finding is consistent with similar research conducted in Mizan-Aman town in South-west Ethiopia^22^ and almost 10 times more likely to utilize PCC services in the cross-section study conducted in West Guji, Ethiopia. ^12^ This result might be due to having a positive attitude toward PCC increases perceived benefit which encourages the individual to use the PCC services. This study shows respondents who had heard about PCC were more than 3 times more likely to use PCC services compared with those who had not heard about PCC AOR=3.15; CI (1.77-5.59). These findings is higher in research conducted in Malaysia and Southeastern Ethiopia showing more than 5 times increase the chances to use PCC services than their counterparts.^21,23^. The possible explanation for this finding is that might be access to information through various sources such as social media, health worker.

This study revealed that the respondents who reach health facilities within 30 minutes are 2.40 times more likely to utilize PCC than those who take more than 30 minutes. This study found similar results with the result conducted in Mizan Aman town, Southwest Ethiopia and the southern part of Ethiopia^22,24^.The availability of health facilities nearer to respondent residential increases the likelihood of utilizing the PCC services. Similarly, participants who had health insurance before pregnancy are 2.68 times more likely to use PCC compared to those without health insurance. The study conducted in Iran shows that there is a relationship between the utilization of PCC services and women having Health insurance (P <0.001). Similarly, the cross-sectional study among 1,368 Jordanian women who had good utilization of PCC who have enrolled in private health insurance (19%) and Government 52.7%)^25,26^.For this the justification might be health insurance reduces the financial burden of health care services thus increasing the chances of seeking the care. Respondents who wait more than 60 minutes at health facilities are 1.60 times more likely to utilize PCC compared to those who wait 60 minutes or less. The possible explanation for this result might be due to more emphasis and value given to health care services.

## Conclusion

The study concludes that in Pokhara Metropolitan there is a low utilization of preconception care services among reproductive-age women. Only 0.3% of participants utilize all preconception care components before their last pregnancy. Only 44.5% had an acceptance level of utilization. The key factors associated with the utilization of PCC services are the age of the respondent, religion, occupation, knowledge, attitude towards PCC, heard about PCC, minutes to reach health facilities, health insurance use before pregnancy, and waiting time at health facilities. These studies highlight the need for targeted interventions to improve awareness and change in attitude towards PCC help to increase PCC services utilization thereby enhancing maternal and child health outcomes.

### Recommendation

- Organize community-based educational programs focusing on the importance of PCC involving local leaders, teachers’ community health workers and community volunteers.
- Use local media to share success stories and positive outcomes of PCC utilization. This can include radio programs and social media campaigns.
- Advocate for health insurance.
- Specific support and necessary counselling given by health facilities to address women with a history of adverse health outcomes.
- Initiation of an appointment system to reduce waiting time at health facilities.

## Data Availability

All relevant data are within the manuscript and its Supporting Information files.

## Acknowledgements

This study was supported by the Health Office, Kaski. The author would like to thank all the participants who gave their valuable time to make this study successful. Most importantly I like to thank Pokhara Metropolitan for their cooperation and assistance in enabling access to the study sites and participants. I also want to acknowledge the School of Health and Allied Sciences, Pokhara University for the technical arrangements and support.

## Author Contributions

Conceptualization and Visualization: Sita Aryal, Dr. Tulsi Ram Bhandari

Data curation: Sita Aryal, Dr. Tulsi Ram Bhandari, Yamuna Marasini

Formal analysis: Sita Aryal, Dr. Tulsi Ram Bhandari, Yamuna Marasini

Investigation: Sita Aryal, Yamuna Marasini, Saroj Wagle

Methodology: Dr. Tulsi Ram Bhandari, Yamuna Marasini

Software: Sita Aryal, Yamuna Marasini

Supervision: Dr. Tulsi Ram Bhandari

Writing– original draft: Sita Aryal, Saroj Wagle

Writing– review & editing: Sita Aryal, Dr. Tulsi Ram Bhandari, Saroj Wagle

All the authors read, revised, and approved the final manuscript.

## Notes

### Competing Interest Statement

The authors have declared no competing interest.

### Funding Statement

The author(s) received no specific funding for this work.

### Author Declarations

Ethical approval was taken from the Institutional Review Committee (IRC) of Pokhara University (Ref No.194-2080/81), before conducting the study.

